# Effect of Pre-Exercise Caffeine Intake on Endurance Performance and Core Temperature Regulation During Exercise in the Heat: A Systematic Review with Meta-Analysis

**DOI:** 10.1101/2021.09.15.21263601

**Authors:** Catherine Naulleau, David Jeker, Timothée Pancrate, Pascale Claveau, Thomas A. Deshayes, Louise M. Burke, Eric D.B. Goulet

## Abstract

**Background:** Heat is associated with physiological strain and endurance performance (EP) impairments. Studies have investigated the impact of caffeine intake upon EP and core temperature (C_T_) in the heat, but results are conflicting. There is a need to systematically determine the impact of pre-exercise caffeine intake in the heat.

**Objective:** Use a meta-analytical approach to determine the effect of pre-exercise caffeine intake on EP and C_T_ in the heat.

**Design:** Systematic review with meta-analysis.

**Data sources:** Four databases and cross-referencing.

**Data analysis:** Weighted mean effect summaries using random-effects models for EP and C_T_, as well as meta regressions with robust standard errors to explore confounders.

**Study selection:** Placebo-controlled, randomized studies in adults (≥ 18 yrs old) with caffeine intake at least 30 min before endurance exercise ≥ 30 min, performed in ambient conditions ≥ 27°C.

**Results:** Respectively 6 and 12 studies examined caffeine’s impact on EP and C_T_, representing 52 and 205 endurance-trained individuals. On average, 6 mg/kg body mass of caffeine were taken 1 h before exercises of ∼ 70 min conducted at 34°C and 47% relative humidity. Caffeine supplementation improved EP by 2.0 ± 0.7% (95% CI: 0.6 to 3.5%) and increased the rate of change in C_T_ by 0.10 ± 0.04°C/h (95% CI: 0.03 to 0.16°C/h), compared with the ingestion of a placebo.

**Conclusion:** Caffeine ingestion of 6 mg/kg body mass ∼ 1 h before an exercise in the heat provides a worthwhile improvement in EP of 2%, while trivially increasing the rate of change in C_T_ by 0.10°C/h.

## 1 Introduction

Many athletes use performance-enhancing supplements during competitive events with the hope of gaining an edge over their opponents. Caffeine, classified as a methylxanthine [1], is a widely used ergogenic aid with robust evidence of performance benefits for a range of exercise and sporting activities [2]. Indeed, since its removal from the World Anti-Doping Agency list of banned substance in 2004 and reclassification to the monitoring list [3], measurement of the caffeine content in urine samples collected at Doping Control during national and international competitions suggested that about 76% of athletes from various sports consumed caffeine prior to or during sporting events [3]. In addition, 89% of athletes competing at the 2005 Ironman™ triathlon world championships revealed that they were planning on using a caffeine substance before and/or during the event [4]. This is not surprising, as pre-exercise caffeine intake of 2-6 mg/kg body mass has generally been shown to confer a worthwhile improvement in endurance performance (EP) under thermoneutral conditions [2, 5-9]. It is proposed that caffeine may contribute to enhance exercise performance by increasing motivation [10] and alertness [11], reducing perceived exertion [12-14] and fatigue [14, 15], enhancing the mobilization of intracellular calcium and free fatty acids [16], and most importantly by acting as an adenosine receptor antagonist [17].

Exercising under warm or hot conditions imposes unique physiological challenges which are ultimately evidenced by the inability of athletes to perform as well as in thermoneutral conditions [18, 19]. Because the extent of changes in physiology [18] and EP [19] during exercise may be dependent upon ambient temperature, humidity and airflow, an honest and precise evaluation of the impact of any supplement that may change physiology on EP can only be reliably obtained while considering the effect of the environment.

The impact of caffeine on EP, C_T_ and physiological functions have been examined with a timing of supplementation occurring either before exercise only, during exercise only or before and during exercise [20]. Peak caffeine concentration in the blood after oral ingestion occurs 30 to 90 min post consumption for low, moderate and high caffeine doses [21]. Moreover, caffeine’s half-life is ∼3-7 h in adults [22]. Hence, any attempt to clearly understand caffeine’s effect on the body during exercise in relation to the dosage administrated is likely to be compromised if more than one caffeine dose is provided. On the other hand, supplementation of a single dose of caffeine before exercise onset provides the opportunity for the compound to produce a predictable and prolonged effect on the metabolism, thereby allowing to better pinpoint, isolate and understand the impact of caffeine during a subsequent exercise.

Several studies have examined the effect of pre-exercise caffeine supplementation on performance during endurance exercise conducted under warm or hot ambient conditions. Overall, results are equivocal, with some showing statistically significant improvements in EP [12, 13, 23, 24] while others did not [14, 25-31]. On the other hand, results of some studies suggest that caffeine supplementation may increase resting energy expenditure [32, 33], diuresis [34, 35], oxygen consumption [36], decrease sweat rate [35] and *ad libitum* fluid intake [35] and impair cutaneous blood flow [37], all of which singularly or in combination may contribute to increase heat stress during exercise. Because of the potential role played by heat strain on performance deterioration [18], several investigations have therefore examined the impact of pre-exercise caffeine intake on core body temperature (C_T_) regulation during exercise [12-14, 23-25, 27-29, 33-35, 38-42]. However, the outcome of this literature is inconclusive, with divergent results regarding caffeine’s impact on EP and C_T_ during exercise being potentially explained by methodological issues, sample sizes and different pre-exercise doses.

Peel, McNarry [43] concluded in a recent meta-analysis that the use of caffeine during exercise in the heat increases C_T_ without providing any ergogenic benefit. While this meta-analysis shed some light on the effectiveness of caffeine ingestion upon EP and C_T_ regulation in the heat, this work only qualified the importance of these effects. Specifically, it did no attempt to determine the magnitude of the changes in EP or C_T_ regulation, which proves to be key and pivotal information for the decision to use or not use caffeine when considering one’s tolerance to heat and the day-to-day normal fluctuation in performance. Importantly, results of studies that provided caffeine before exercise onset were combined with those where caffeine was taken both before and during exercise (i.e., maintenance dose), which may have confounded the results.

The combination of the increasing number of sporting events taking place in warm or hot environments and the popularity of caffeine as an ergogenic aid provide a strong rationale for a specific and thorough review of the effect of caffeine supplementation on EP and C_T_ regulation during exercise performed under warm or hot conditions. Therefore, this meta-analysis aims to assess the magnitude of the effect of pre-exercise caffeine intake on EP (1) and C_T_ regulation in warm or hot conditions (2) and to identify potential factors that may impact the relationship between the control of C_T_ and caffeine intake (3). The results of this meta-analysis will be useful for athletes and their support teams to evaluate the benefits and risks of an acute caffeine consumption before endurance events in warm or hot conditions.

## 2 Methods

### 2.1. Search strategy for identification of studies

The search strategy used for study selection is presented in Figure 1. A first search was performed using cross-referencing of the reference sections of five meta-analyses [2, 5, 7, 44, 45] retrieved in an umbrella meta-analysis [8] and two narrative reviews [9, 46]. Then, a traditional literature search, limited to original peer-reviewed articles published in French or English, was performed on MEDLINE, SPORTDiscus, AMED and CINAHL databases, combining a “title field” and an “abstract field” research using the following keywords alone or in combination with truncation: caffein*, dehydrat*, endurance, exercise, heat, hot, humid*, temper*, therm*, performance. The exact search strategy can be found in supplementary material 1. The last search of the literature was done on September 1^st^ 2021.

**Figure 1.**
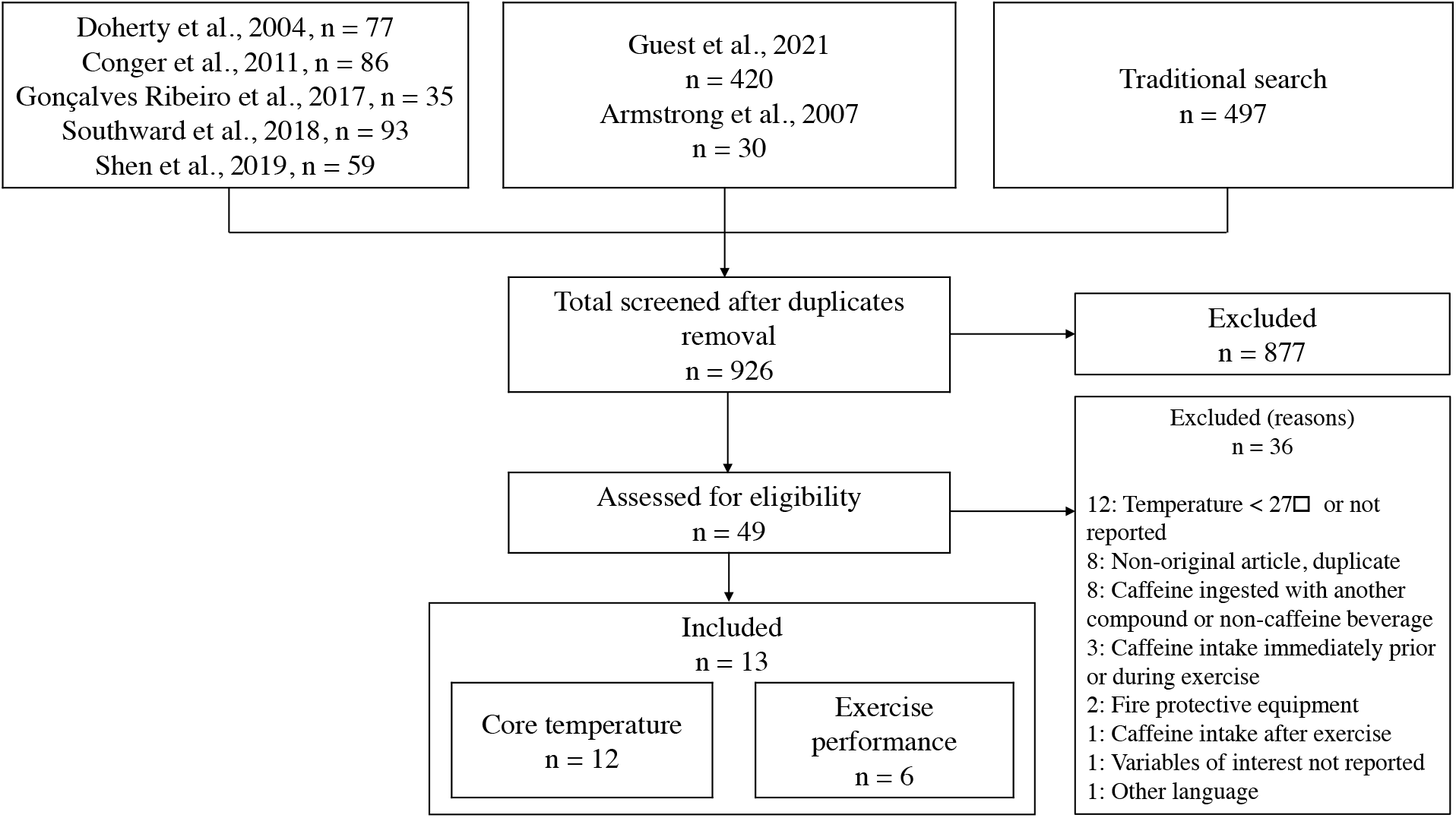
Flowchart showing the selection process used for the inclusion and exclusion of studies.

All references were merged in an Excel spreadsheet and duplicates were removed. Then, a first selection based on the title was performed; afterward, the abstract and method sections of all potential articles were read. When exercise was performed in hot or warm conditions, caffeine was ingested and EP or C_T_ measured, the methodological section was read to verify eligibility. Thesis, published abstracts, case studies, non-peer-review manuscripts and conference proceedings were not considered. When needed, authors of included studies were contacted and asked to share raw data or provide further details/missing values.

### 2.2. Criteria for considering studies for inclusion in the meta-analysis

Inclusion criteria were: (1) placebo-controlled, randomized studies on healthy adults ≥ 18 yrs old; (2) endurance exercise including cycling, running and walking ≥ 30 min; (3) ambient temperature ≥ 27°C; (4) caffeine intake at least 30 min before exercise and no caffeine provided during exercise and; (5) C_T_ measured as rectal, esophageal or gastrointestinal temperature. Studies were excluded when: (1) caffeine was ingested with another compound, with the exception of water; (2) caffeine was not swallowed (i.e., mouth rinse); (3) participants wore fire protective equipment which would affect thermoregulation and; (4) intermittent exercise was performed.

### 2.3. Assessment of trial quality

Trial quality assessment using a scale can influence the interpretation of results of meta-analyses [47]; thereby, trial quality assessment was not performed in the present meta-analysis.

### 2.4. Data extraction

Data regarding characteristics of the (1) studies; (2) participants; (3) exercise protocols; (4) caffeine intake protocols and; (5) EP or C_T_ were extracted and coded in spreadsheets using double data entry. Any disagreement was discussed, and a consensus was reached. When not provided by authors, data only available in figures were extracted using WebPlotDigitizer.

### 2.5. Confounding variables

The effect of *a priori* identified potential confounding variables on the relationship between caffeine intake and EP or C_T_ such as ambient temperature, humidity, exercise duration, exercise intensity, caffeine dose and the elapsed time between caffeine ingestion and the onset of exercise was verified. For Cohen, Nelson [26], only wet-bulb globe temperature was reported. Therefore, ambient temperature and humidity were taken as found in Armstrong, Casa [46].

### 2.6. Exercise duration and intensity and participants 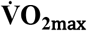

Total exercise duration was computed as the average exercise time completed during both the caffeine (experimental) and placebo conditions. When a pre-load was performed prior to a performance test, it was included in the total exercise duration time. Exercise intensity was taken as the average of the mean % maximal oxygen consumption 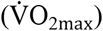 at which both conditions were performed. A weighted average technique was used for studies that utilized a combination of exercise intensities [48]. When not measured by authors, exercise intensity was estimated and computed as explained by Goulet [48] and Goulet [49]. Finally, for Cohen, Nelson [26], exercise intensity could not be calculated due to a lack of data.

### 2.7. Assessment of endurance performance

To assist the understanding and interpretation of findings, all performance outcomes were transformed and are reported as % changes in power output between the caffeine and placebo conditions. For studies that used a time-trial type exercise protocol and reported the mean power output during exercise, the percent changes in EP were calculated using the following equation: Mean power output in the caffeine condition (W) - mean power output in the placebo condition (W) / mean power output in placebo condition (W) × 100 (1)

Endurance performance reported in kJ was converted to power output (W) [12, 25]. The same formula was used for studies that reported time-trial completion time, but results were multiplied by -1 so that a reduction in time, was converted to a positive value to indicate an increase in performance. For Ping, Keong [13] that used a fixed-intensity test to exhaustion, performance was transformed to percent changes in power output (W) using the following equation [50]: Mean time to exhaustion (min) in the caffeine condition - mean time to exhaustion (min) in the placebo condition / mean time to exhaustion (min) in the placebo condition × 100 / 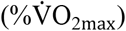 at which the test was performed / 6.4) (2)

Conversions between time trial and power outputs were undertaken with the assumption that a 1% change in running time-trial time equals a 1% change in power output, and that a 0.4% change in cycling time-trial time equals a 1% change in power output [50].

Out of the 6 [12, 13, 25-27, 29] studies that met the inclusion criteria for EP, access to raw experimental data was available for 4 investigations [12, 25, 27, 29]. Accordingly, for these studies, mean changes in EP and associated standard errors were computed directly from available raw data. The remaining two studies [13, 26] provided the mean EP data obtained with the placebo and caffeine conditions. For Ping, Keong [13] the variance associated with the change in EP was calculated from the provided *p*-value, assuming a value ≤ 0.05 to represent *p* = 0.05. Cohen, Nelson [26] provided no variance associated with the mean change in EP. It was therefore estimated using an imputed correlation coefficient of 0.88, representing the weighted mean correlation coefficient computed from values of 4 [12, 25, 27, 29] individual studies.

### 2.8. Assessment of core temperature changes during exercise

Changes in C_T_ were assessed using from measurements taken from esophageal [39, 42], gastrointestinal [12, 27] or rectal [13, 23-25, 29, 34, 35, 38] regions; these areas provide slightly different figures of C_T_ during exercise. However, this is irrelevant in the context of the current meta-analysis where we were focused on changes in C_T_ within each of the included studies. More specifically, the variable of interest was taken as the rate (°C/h) of change in C_T_ within each of the placebo and caffeine conditions of each of the included studies. When more than two assessments of C_T_ were performed within a single study, the mean rate of change in C_T_ from one measurement point to the other within a condition was computed for each of the placebo and caffeine condition. To obtain the overall mean rate of change in C_T_ between the placebo and caffeine conditions within a single study, first, the mean rate of change in C_T_ observed between measurement points within the placebo condition was subtracted from that obtained within the caffeine condition and, second, the mean and associated standard error of all changes were computed. Ping, Keong [13] measured C_T_ only at the start and end of exercise for both the caffeine and placebo conditions. For this study, first, the mean rate of change in C_T_ in the placebo and caffeine conditions was computed by subtracting the first from the last measurement of C_T_ and, second, using the *p-*value associated with the change in C_T_ within each of the conditions the standard errors were computed. To obtain the overall mean rate of change in C_T_ between the placebo and caffeine conditions, the mean rate of change in C_T_ observed with the placebo condition was subtracted from that obtained in the caffeine condition, and the associated standard error was computed using a correlation coefficient between measurements of 0.50 [51].

### 2.9. Statistical analyses

#### 2.9.1. Software

Data were analyzed using the Microsoft Office Excel 2020 (version 1902, Redmond, WA, USA), MetaXL [52], Comprehensive Meta-Analysis (version 2.2.064, Englewood, NJ, USA), STATA/MP (version 14, College Station, TX, USA) and IBM SPSS Statistics (version 21, Armonk, NY, USA) software.

### 2.10. Weighted mean effect summaries

The weighted mean effect summaries were determined using method of moment random-effects model, and results were considered statistically significant when the confidence intervals (CI) did not include 0. Some studies examined the effect of more than one caffeine dose on EP or C_T_ regulation during exercise. To prevent the violation of the assumption of independence among research observations, data which examined the effect of different caffeine doses within the same study were merged [24, 26, 27], considering the caffeine dose to be representative of the average dose provided. Del Coso, Estevez [34] provided a single dose of caffeine but performed experiments where the placebo and caffeine were provided with or without water; for both the placebo and caffeine conditions results of those two trials were combined. Hunt, Hospers [42] examined the impact of caffeine consumption in habituated and non-habituated consumers; for this study, data from both trials were merged. Studies examined C_T_ regulation during fixed-intensity as well as during time-trial conditions. We verified whether there was a difference in the rate of increase in C_T_ between the different exercise protocols; the difference was < 0.1°C/h. Therefore, data from both exercise protocols were aggregated for analyses. Results of the weighted mean effect summaries are reported as means ± standard errors and/or 95% CI.

### 2.11. Practical significance of the weighted mean effect summaries

The qualitative interpretation of the practical significance of the effect of caffeine supplementation on EP and C_T_ regulation during exercise were performed as recommended by Hopkins [53], while using minimal threshold changes in EP and C_T_ of respectively 1.5% [54] and 0.25°C [55].

### 2.12. Heterogeneity, publication bias and sensitivity analysis

Cochran’s Q and I^2^ statistics were both used to assess between-study heterogeneity and the degree of inconsistency among results of included studies [56]. Cochran’s Q test was considered significant at *p* ≤ 0.1 [57]. The following classification was used to interpret the I^2^ statistic: low (< 40%), moderate (40-59%), substantial (> 60%) [58]. Publication bias was performed using visual assessment of funnel plots with Trim and Fill adjustments [59]. A sensitivity analysis was performed on each of the forest plots by removing each study once from the models to determine whether this would change the magnitude of the outcome summaries.

### 2.13. Meta-regression analyses

Meta-regressions were performed by regressing the mean differences in the rate of increase in C_T_ between the placebo and caffeine conditions on each of the *a priori* identified confounders; no regression analyses were performed regarding EP due to the low number of included studies. Meta-regression analyses were performed using method of moment random-effects model, with 95% robust (Huber-Eicher-White-sandwich) standard errors [60].

## 3 Results

### 3.1. Search results and characteristics of the included studies

After duplicates were removed, a total of 926 titles were evaluated (Figure 1). In the remaining 49 articles assessed for eligibility, a total of 13 [12, 13, 23-27, 29, 34, 35, 38, 39, 42] were included in the meta-analysis. Six of these investigations [12, 13, 25-27, 29] provided data related to EP, 12 papers [12, 13, 23-27, 29, 34, 35, 38, 42] reported on C_T_ regulation during exercise (Table 1), and 5 studies [12, 13, 25, 27, 29] contained both EP and C_T_ data. The studies were published between 1994 and 2021 in 9 different peer-reviewed journals. Seven studies were conducted in the USA [23-27, 35, 38] and 1 in each of those countries: Australia [42], Belgium [29], Japan [39], Malaysia [13], Spain [34] and the UK [12].

**Table 1.**
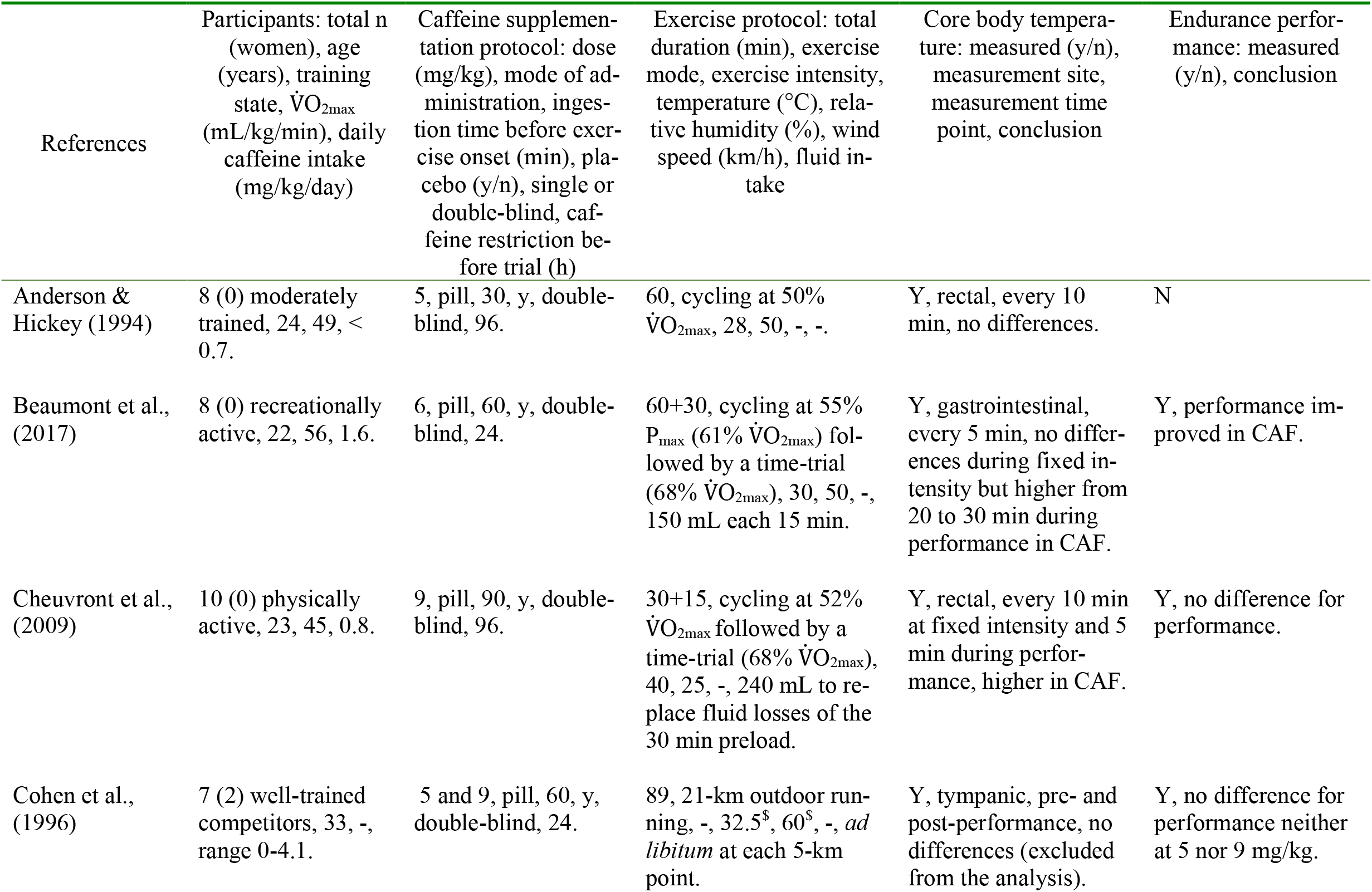
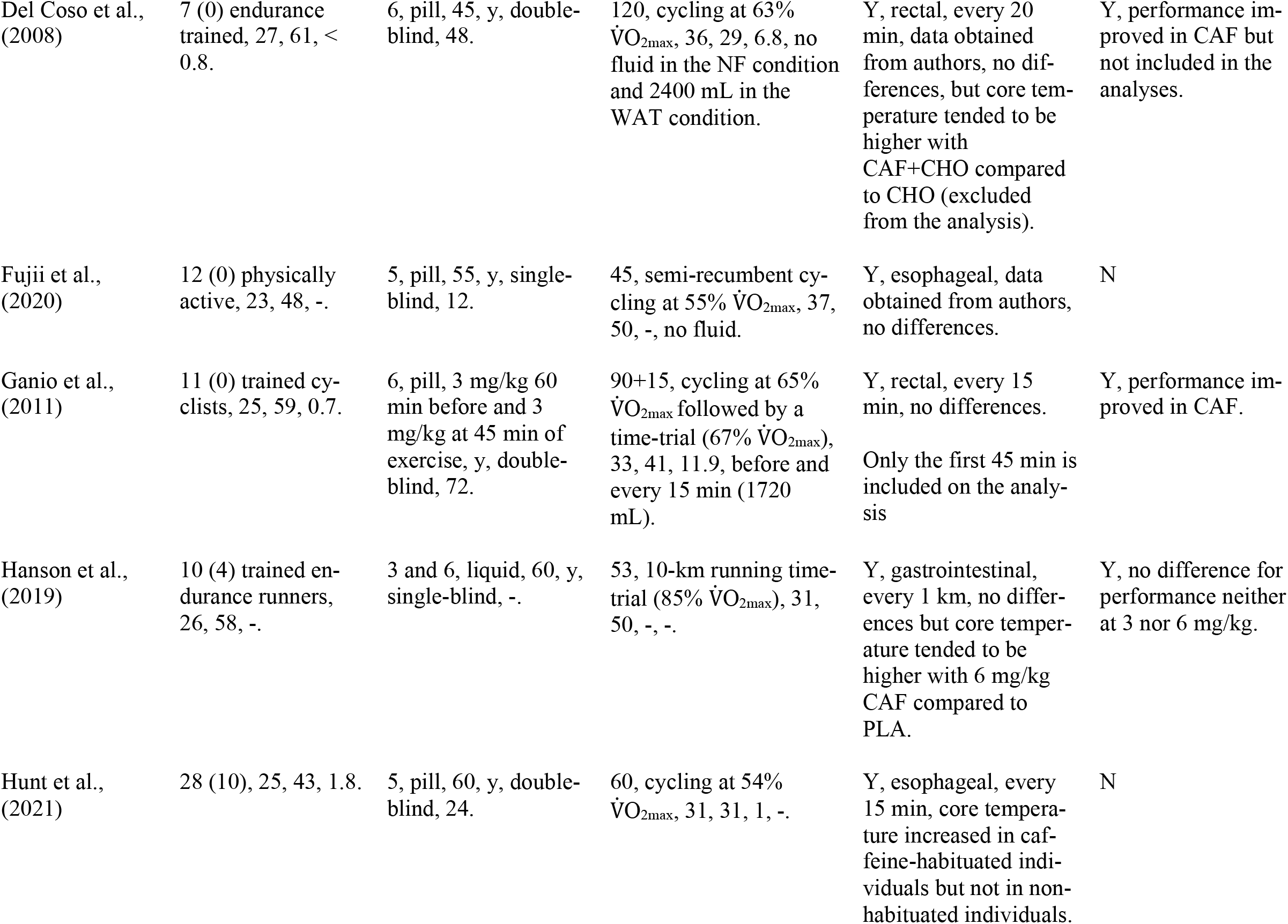
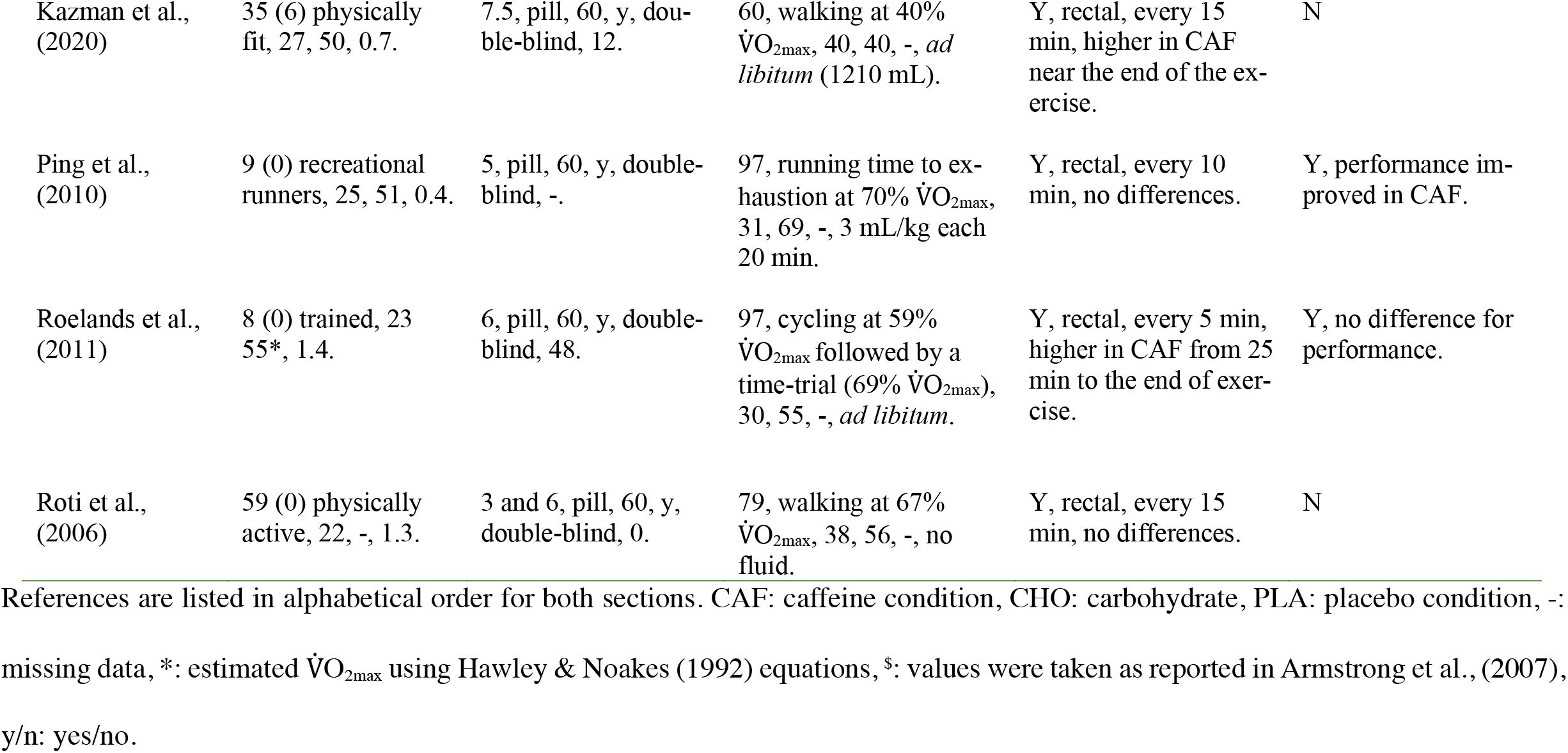
Summary of characteristics of included studies.

### 3.2. Participants’ characteristics

A total of 212 physically active to well-trained individuals was represented among the 13 studies included in the meta-analysis, with women representing 10% of the total participants. Mean sample size was 16 ± 15 individuals per study (range 7-59). More specifically, results for EP were derived from a total of 52 participants, whereas those for C_T_ regulation were provided by 205 participants, with women representing 12 and 10% of the subsamples, respectively. The mean age, height, body mass and 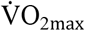 of the participants for those studies which assessed EP and C_T_ regulation were respectively 25 ± 4/24 ± 2 yrs, 176 ± 5/176 ± 4 cm, 71 ± 8/73 ± 6 kg and 53 ± 5 (n = 5)/52 ± 6 (n = 11) mL/kg/min.

### 3.3. Characteristics of the placebo/caffeine administration and exercise protocols

All studies included placebos and were conducted using either single [27, 39]- or double [12, 13, 23-26, 29, 34, 35, 38, 42]-blinded fashion. With the exception of Roti, Casa [24], all studies were performed in a crossover manner where participants acted as their own control. The mean characteristics for the protocols used in the studies that assessed EP and C_T_ regulation respectively were caffeine dose (6 ± 2; 6 ± 2 mg/kg body mass), elapsed time between the placebo or caffeine intake and onset of exercise (65 ± 12; 58 ± 14 min), caffeine restriction time leading to the experiments (48 ± 34 (n = 4); 43 ± 35 h (n = 10)), ambient temperature and relative humidity (32 ± 4, 34 ± 4°C; 52 ± 15, 46 ± 13%), exercise duration (79 ± 23; 70 ± 24 min) and exercise intensity 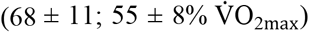. Wind speed was reported in only three studies [23, 34, 42]; therefore, these values are not presented. Exercise protocols consisted of either a running time-trial [26, 27], a fixed-intensity cycling or walking period [24, 34, 35, 38, 39, 42], a pre-load cycling period followed by a time-trial [12, 23, 25, 29] or a running test to exhaustion [13].

### 3.4. Weighted mean effect summaries

Except for one study that demonstrated a negative impact of caffeine supplementation on EP [29], caffeine intake prior to exercise improved performance in all other studies, with gains ranging in magnitude from trivial to important (Figure 2). The combined results showed a performance improvement with pre-exercise caffeine supplementation of 2.0 ± 0.7% (95% CI: 0.6 to 3.5%), with an 80% chance to confer a worthwhile improvement in cycling and running performance under field conditions. Removing each study once from our model revealed changes in EP fluctuating between 1.4 to 2.4%. Distribution of point estimates around the weighted mean effect summaries was appropriate, which suggests no publication bias. Heterogeneity was low with an *I*^2^ of 26% and a Cochran’s *Q* of 6.8, *p =* 0.2.

**Figure 2.**
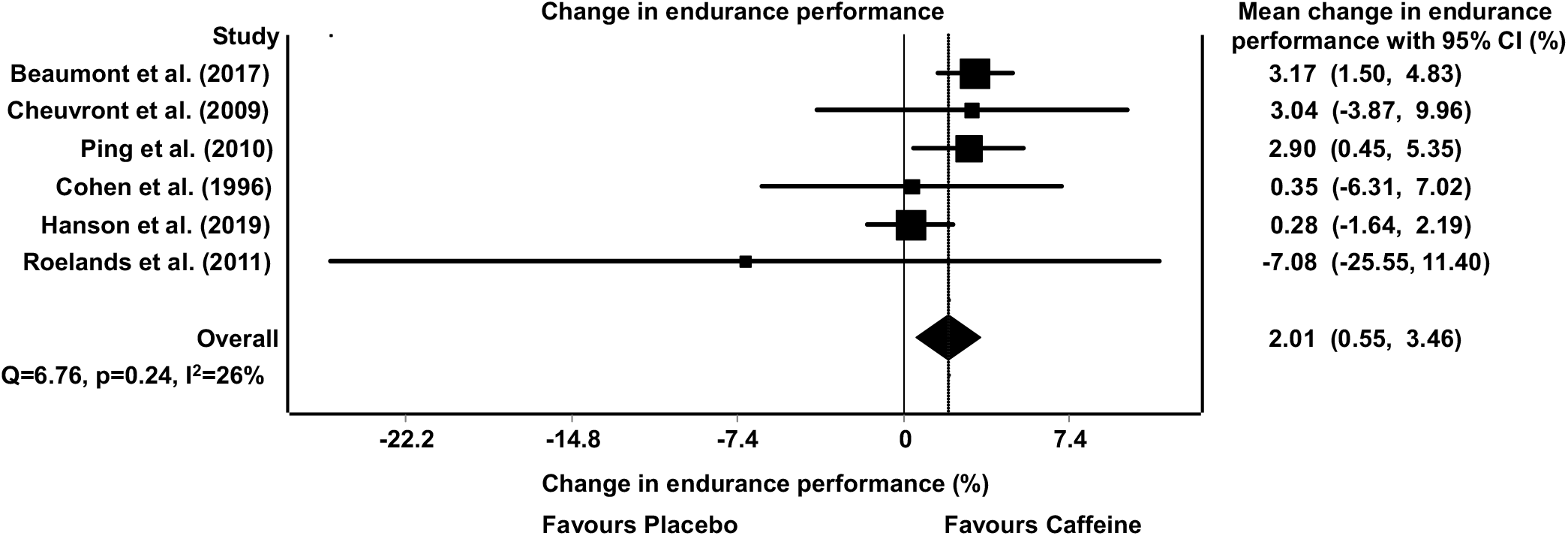
Forest plot showing the mean weighted change in endurance performance between the caffeine and placebo conditions. CI: confidence interval.

Figure 3 shows that caffeine supplementation increased the rate of change in C_T_ by 0.10 ± 0.04°C/h (95% CI: 0.03 to 0.16°C/h), compared with the ingestion of a placebo which, from a practical perspective, represents a trivial change over a 60 min period of exercise. None of the studies included in the model affected the rate of change in C_T_ more than any other study. Indeed, reanalysis of findings with the exclusion of any one study once from the model revealed changes in the rates of increase in C_T_ varying from 0.07 to 0.12°C/h. Examination of the funnel plot shows that there was an appearance of a slight publication bias with more studies at the bottom right than left of the mean. However, a trim and fill analysis indicated that adjusting for those missing studies decreases the mean rate of change in C_T_ from 0.10 to 0.07°C/h, which represents a trivial adjustment in the mean, but would nevertheless reverse the statistical significance of the weighted mean effect summary with a 95% CI of -0.002 to 0.148. Inconsistency among research observations was moderate with an *I*^2^ of 36% and a Cochran’s *Q* of 17.1, *p* = 0.10. Studies that included time-trials (0.07 ± 0.03, 95% CI: 0.02-0.12, n = 5) in their exercise protocol did not show a greater rate of increase in C_T_ compared to studies using only fixed-intensity exercise (0.13 ± 0.07, 95% CI: 0.006-0.040, n = 7)

**Figure 3.**
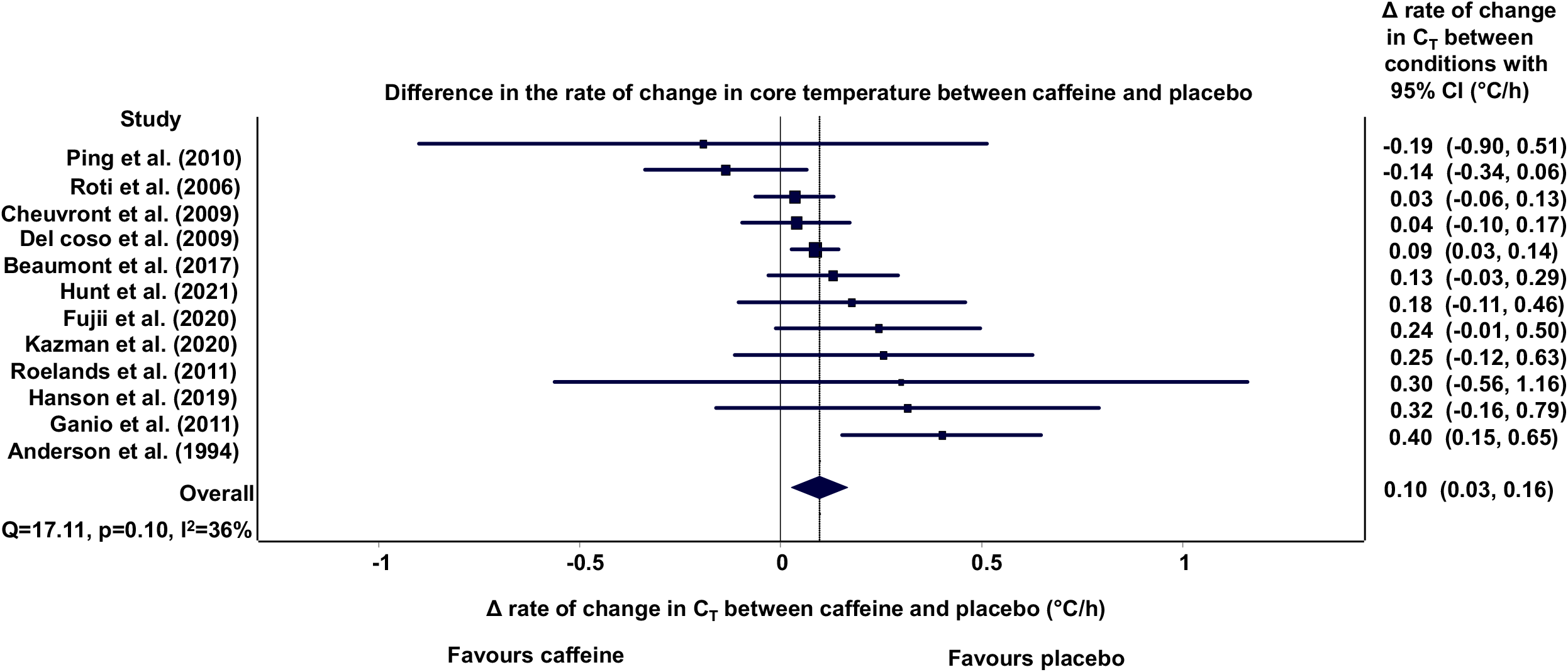
Forest plot showing the mean weighted difference in the rate of increase in core temperature between the caffeine and placebo conditions. CI: confidence interval; C_T_: core temperature. Δrate: caffeine - placebo. Favours caffeine: rate of increase in core temperature is lower with caffeine; Favours placebo: rate of increase in core temperature is lower with placebo.

### 3.5. Meta-regression analyses

Figure 4 demonstrates that there was no significant relationship between the changes in the rate of increase in C_T_ between the caffeine and placebo conditions during exercise and the caffeine dosage (a) or the timing between caffeine ingestion and the start of exercise (b). Figure 5 shows the correlations between the changes in the rate of increase in C_T_ in the caffeine and placebo conditions during exercise and temperature (a), relative humidity (b), exercise duration (c) and exercise intensity (d). With the exception of exercise intensity, none of the slopes was statistically significant. Except for relative humidity, results highlighted the fact that as temperature, exercise duration and exercise intensity increased, there was a progressive decrease in the rate of C_T_ rise with caffeine, compared with the placebo. The model revealed that at a threshold ambient temperature of 42.3°C and exercise intensity of 68% of 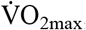, the rate of the rise in C_T_ with caffeine should be lower than with the placebo. Combining exercise intensity and temperature in the same model explained as much as 61% of the rise in C_T_, with relative humidity, exercise duration and caffeine dose contributing trivially to improve the precision of the model (∼1%).

**Figure 4.**
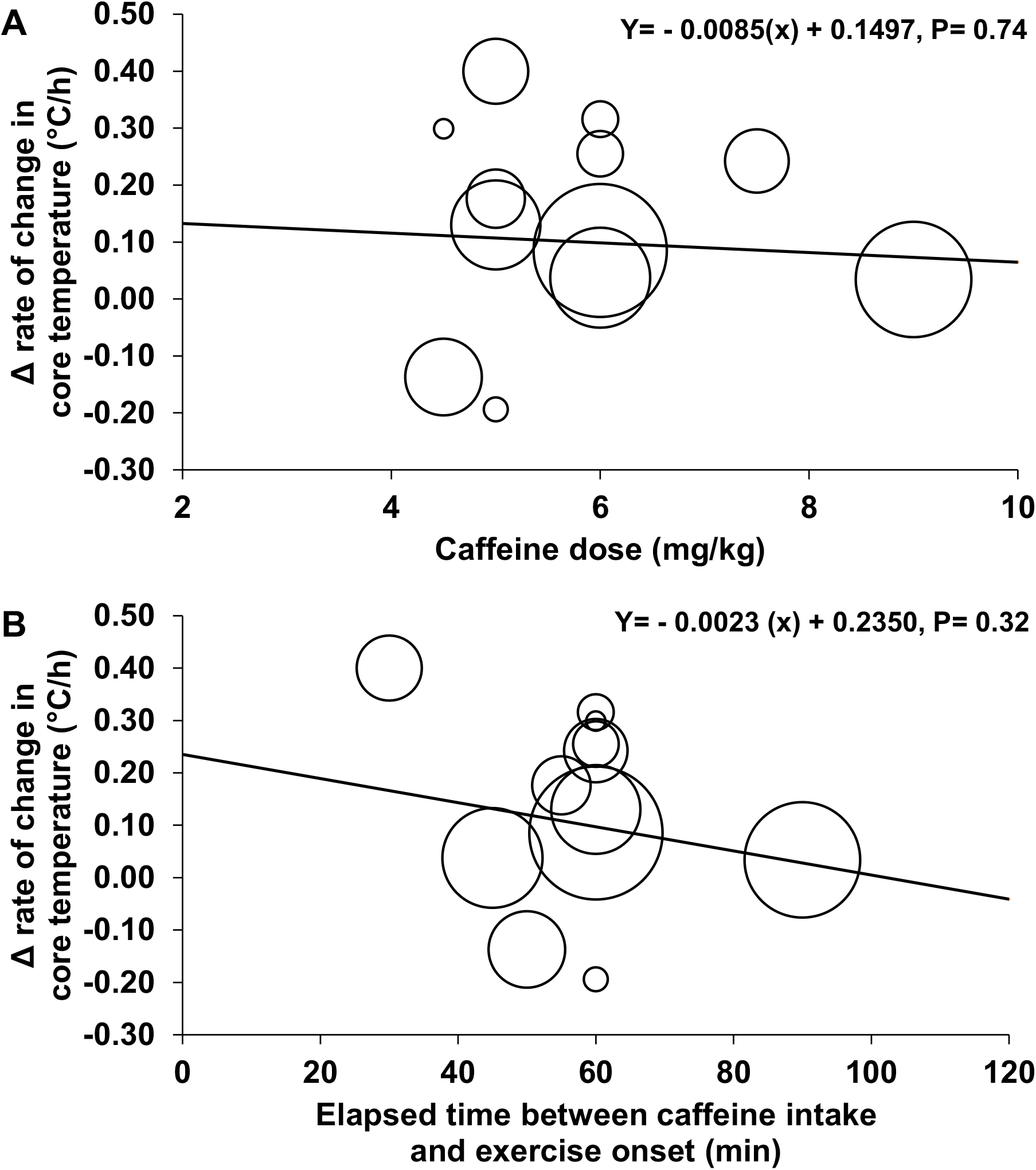
Relationships between the changes in the rate of increase in core temperature between the caffeine and placebo conditions and caffeine dose (a) and elapsed time between caffeine intake and exercise onset (b). Δrate: caffeine - placebo.

**Figure 5.**
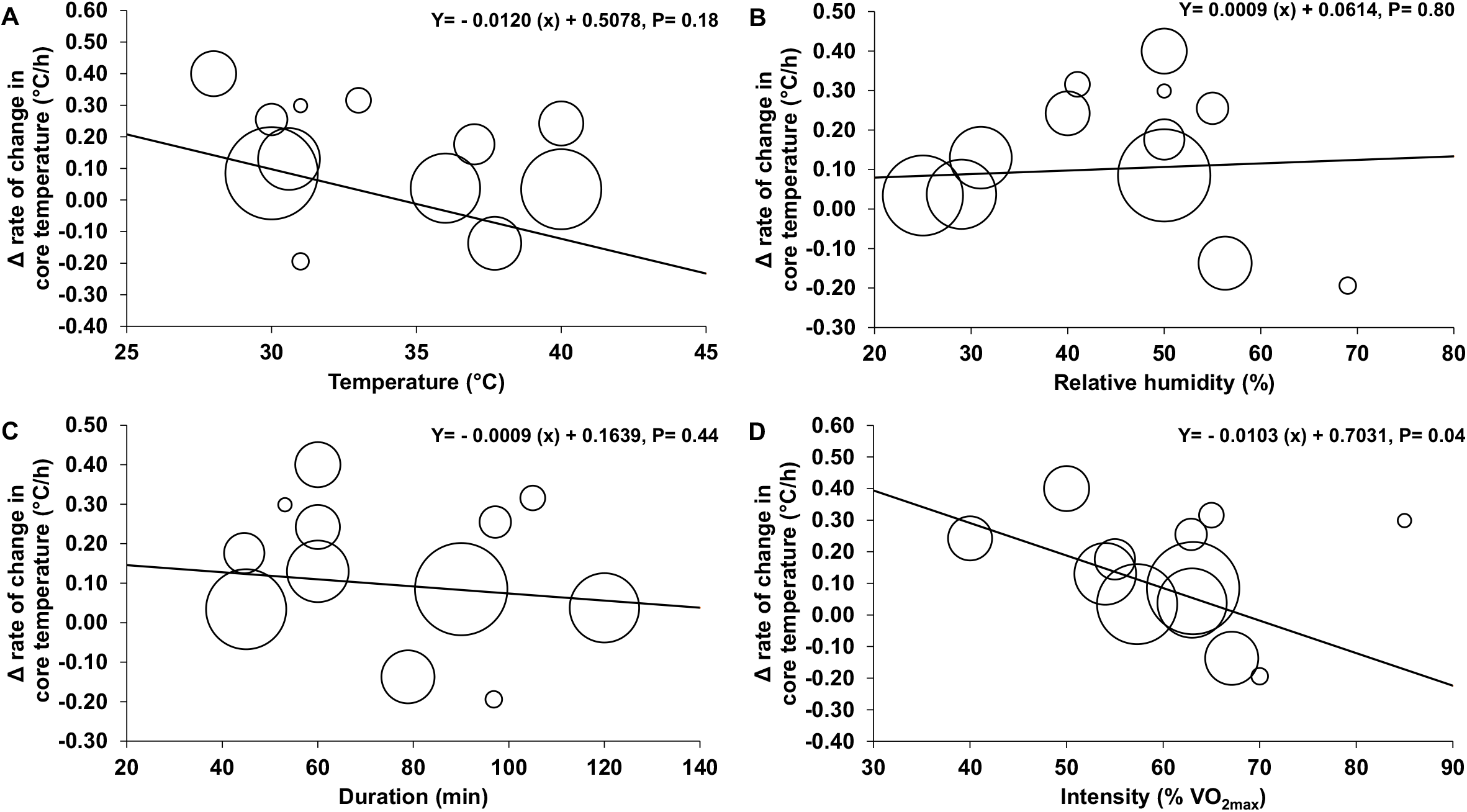
Relationships between the changes in the rate of increase in core temperature between the caffeine and placebo conditions and temperature (a), relative humidity (b), exercise duration (c) and exercise intensity (d). Δrate: caffeine - placebo.

## 4. Discussion

While several systematic reviews, including meta-analyses [2, 5, 7, 44, 45] and an umbrella review [8], have reported on the impact of pre-exercise caffeine ingestion on EP, specific investigations of its impact in warm or hot environments are limited. Therefore, the goal of the current work was to extend that of the previous meta-analyses by exclusively focussing on the impact of pre-exercise caffeine ingestion on EP and C_T_ during exercise undertaken in the heat. Our results indicated that caffeine (1) provides a statistically significant and worthwhile improvement in EP and (2) is associated with a significant, although practically trivial, increase in the rate of rise in C_T_ in warm or hot conditions.

Our results show that pre-exercise caffeine supplementation improves EP in a practically meaningful manner, which contradicts with the findings of Peel, McNarry [43] who observed a trivial effect of caffeine supplementation on EP in a warm environment. The discrepancy between findings might, on the one hand, be linked to the criteria used for study inclusion. Indeed, while we limited our analysis to those studies that provided a single dose of caffeine prior to exercise onset, Peel, McNarry [43] amalgamed all studies conducted in the heat, irrespective of the moment of caffeine administration relative to exercise onset or the number of doses provided during the protocol. On the other, we computed the percent change in performance between the placebo and caffeine trial and compared it to a threshold relative to the day-to-day normal fluctuation in EP. In contrast, Peel, McNarry [43] limited their analysis to the reporting of an Hedge G’s.

The mean EP improvement of 2% observed in the current meta-analysis is similar to the outcome derived from the meta-analysis undertaken by Southward, Rutherfurd-Markwick [2]. Indeed, their assessment of a mix of studies completed in a wide range of ambient temperatures, including heat, identified an improvement in mean power output of 3% associated with caffeine ingestion. However, this investigation lacked a specific analysis of the impact of ambient temperature on this effect. Nevertheless, given the low proportion of studies conducted in the heat in this analysis, which implies that the weight of those performed in a cooler environment was much larger, our results could suggest that pre-exercise caffeine intake has a similar effect on EP in hot and temperate environments.

To test this issue within a single study, Ganio, Johnson [23] investigated a protocol involving 90 min of cycling followed by a 15 min performance trial at 12 or 33°C, in which participants consumed 3 mg/kg body mass of caffeine before and during exercise (total of 6 mg/kg body mass). Compared with the placebo, caffeine intake improved EP in the heat, although not significantly, close to twofold compared with the temperate conditions (4% at 12°C; 7% at 33°C). Whether these findings support our assertion that pre-exercise caffeine intake has a similar effect on EP in temperate and hot environments or, to the contrary, that caffeine intake may confer an advantage in the latter environment is not clear. However, it must be considered that this study provided caffeine during exercise and one of the conditions was conducted in a cool rather than a temperate environment. Nevertheless, as potentially suggested by the results of this studyGanio, Johnson [23], pre-exercise supplementation with caffeine may be particularly relevant for endurance exercise conducted in the heat. Indeed, caffeine enhances dopamine signaling in the brain [61] through its effect on adenosine receptors [62, 63]. In a series of studies conducted in a temperate (18°C) and hot environment (30°C), Watson, Hasegawa [64] observed that the pre-exercise administration of a dopamine/noradrenaline reuptake inhibitor failed to alter time-trial effort in the temperate condition, but was associated with a 9% improvement in the heat. It was proposed that the augmented brain concentration of dopamine counteracted the hyperthermia-induced decrease in motivation and drive, thereby allowing an improved performance [65].

Our results showed that, compared with a placebo, pre-exercise caffeine ingestion significantly increases the rate of change in C_T_ during exercise in warm or hot environments by ∼ 0.10°C/h. However, it is important to underline that, from a practical point of view, such a rate of increase in C_T_ is trivial, at least for short endurance exercise duration of 1-2 h. Indeed, the normal daily fluctuation in core temperature is of the order of ∼ 0.25°C [66]. Why caffeine appears to increase the rate of the rise in C_T_ during exercise is not clear. However, analyses showed that the changes in the rate of increase in C_T_ between conditions do not seem to be related to confounders such as ambient temperature, relative humidity, exercise duration, nor the dosage and the timing of the caffeine intake prior to exercise. Interestingly, at higher exercise intensities, there was a significant decline in the rate of the rise in CT with caffeine compared with placebo; specifically, our modelling showed that at exercise 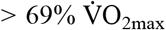, the rate of the rise in CT was greater in the placebo condition. Nonetheless, such results must be interpreted carefully given that only simple meta-regressions were performed and that other confounding factors could have influenced this relationship. Notwithstanding, this observation may provide some reassurance that during a typical training or race, pre-exercise caffeine intake is unlikely to have a significant impact on thermoregulation.

Whether the greater rate of rise in C_T_ with caffeine can be attributed to an enhanced rate of heat production or a lowered rate of heat dissipation is unknown. However, results of previous studies do not support impaired heat loss mechanisms, even in the heat, as a potential concern [23, 33]. Hence, an increase in internal heat production may potentially account for the greater rate of rise in C_T_ with caffeine ingestion. Although speculative, the increased C_T_ with caffeine ingestion may be caused by the higher dopamine levels within the brain. Indeed, it has been demonstrated that intracerebroventricular injection of dopamine produces hyperthermia in rabbits and goats [67]. Several studies that showed that caffeine supplementation increased time-trial EP also demonstrated a greater rate of rise in C_T_, compared with the placebo [12, 25, 27, 29]. In those studies, of course, part of the greater rise in C_T_ with caffeine may simply be explained by the greater intensity of exercise.

Of potential practical importance is the consideration that differences in the rate of the rise in C_T,_ if this remains constant during prolonged exercise (i.e., > 2 h), may expose athletes who participate in ultra endurance events such as Ironman™ or ultra-marathons in hot conditions to higher risks of heat-related fatigue or illnesses after caffeine consumption. This assertion is particularly relevant given the likelihood that such athletes will consume carbohydrates (CHO) and take caffeine maintenance doses throughout the events. Here, maintenance of high dopamine levels in the brain, as a result of recurrent intake of caffeine during exercise, could maintain the drive for the caffeine-induced increase in C_T_. Furthermore, close observation of pertinent literature suggests that the combination of caffeine and CHO intake during exercise may interact to increase C_T_. For instance, compared with the intake of CHO alone during exercise, several studies [31, 34, 68] have observed an increased C_T_ when caffeine was provided before exercise along with CHO + caffeine during steady-state exercise. It is unclear why the co-ingestion of caffeine and CHO appears to induce a disproportional increase in C_T_. Although speculative, it may be related to the fact that CHO ingestion has been shown to increase dopamine release in the brain, at least in rodents [69]. Moreover, caffeine may facilitate intestinal CHO absorption via an enhanced sodium-glucose-linked transporter activity [70]. An increased absorption of CHO would likely be concomitant with an elevated splanchnic blood flow [71] causing a slight reduction in skin blood flow [72], thereby leading to an impaired thermoregulation.

Several aspects need to be considered and kept in mind when interpreting the current results. First, the literature search was limited to English and French citations; thus, studies published in other languages may have been missed. Our results may not apply to prolonged exercise, i.e., > 2 h. The exercise intensity and level of the athletes taking part in the included studies may not be comparable to real-world competitive endurance events. Finally, the present results only apply to well-trained adults, primarily males, for walking, running and cycling exercises of ∼ 80 min in duration conducted in warm or hot environments with caffeine doses of 3-9 mg/kg taken ∼ 1 h before the onset of exercise.

## 5 Recommendations for future studies

To pursue our understanding of the impact of caffeine ingestion on EP and C_T_ regulation under warm or hot conditions, we propose the following perspectives for future studies. 1) Competitive athletes frequently consume caffeine before and during events. They also compete at high intensities and, often, for longer than 60 min. These factors combined increase metabolic heat production, which could have a large influence on heat storage. Whether this may render those athletes more susceptible to heat-related fatigue or illnesses needs to be determined. 2) Co-ingestion of caffeine and CHO is common in competitive athletes during prolonged exercise and there is a strong possibility that these compounds could interact to disproportionally increase C_T_. There is a need to determine the impact of their combined ingestion on C_T_ during prolonged exercise. 3) Research in female athletes is scarce. Given the effect of the menstrual cycle on thermoregulatory controls [73] there is a need to examine caffeine’s impact on EP and C_T_ during the different phases of the menstrual cycle. 4) Recent work has demonstrated that untrained habituated caffeine consumers show a greater rise in C_T_ with caffeine than a placebo during exercise, which is not the case in non-habituated consumers for whom the increase in C_T_ with or without caffeine is similar. Whether this observation may also apply to competitive individuals needs to be determined. 5) All future studies designed for and conducted in competitive athletes should increase their efforts to replicate real-world ambient conditions to adequately reflect competitive settings, especially ensuring to mimic the effect of convection [74] and radiation [75] as closely as possible.

## 6 Conclusion

Findings of the present meta-analysis showed that caffeine ingestion before endurance exercise lasting ∼ 70 min in warm or hot environments improves performance by 2% and increases C_T_ by 0.10°C/h. The enhancement in EP is considered practically worthwhile, while the impact on thermoregulation is considered trivial. Thus, based on the current results, athletes and their support teams should feel confident in including caffeine as an ergogenic aid strategy before competitive endurance events performed in warm or hot conditions.

## Data Availability

Not available. All of the data used in the present meta-analysis are available in the included orgininal studies.

## Key Points

What is already known?

1. Pre-exercise caffeine ingestion of 2 to 6 mg/kg body mass is known to improve EP in a meaningful way.
2. Several studies have investigated the impact of caffeine ingestion upon EP and C_T_ regulation under warm or hot environments. Results are divergent but, yet, no meta-analysis has been conducted to determine the magnitude of the effect of caffeine supplementation on these parameters.

What are the new findings?

1. From a statistical point of view, EP and the rate of increase in C_T_ are enhanced by pre-exercise caffeine ingestion of 3-9 mg/kg body mass 1 h before exercise onset by 2% and 0.10°C/h, respectively.
2. From a practical point of view, the impact of pre-exercise caffeine ingestion on EP is important, while that on the rate of increase in C_T_ is trivial.

## Data availability statement

All the data presented in the article will be made available from the corresponding author upon reasonable request.

## Acknowledgments

Thomas A. Deshayes is financially supported by the Fonds de Recherche du Québec - Santé (FRQS). The authors wish to thank the researchers who shared experimental data and provided further information.

## Authors’ contributions

CN, DJ, TP, PC, TAD and EDBG: designed the research and performed the literature search. TP, TAD and EDBG performed the data extraction. TAD and EDBG: performed the statistical analyses. TP, TAD and EDBG: designed the tables and figures. CN, DJ, TP, PC, TAD, LMB and EDBG: interpreted data, drafted and revised the manuscript. All authors approved the final version of the manuscript.

